# Stepwise school opening online and off-line and an impact on the epidemiology of COVID-19 in the pediatric population

**DOI:** 10.1101/2020.08.03.20165589

**Authors:** Yoonsun Yoon, Kyung-Ran Kim, Hwanhee Park, So young Kim, Yae-Jean Kim

**Affiliations:** Department of Pediatrics, Samsung Medical Center, Sungkyunkwan University School of Medicine, Seoul, Republic of Korea; Department of Mathematics, Konkuk University, Seoul, Republic of Korea

**Keywords:** COVID-19, children, adolescents, pediatric, school, opening, classes, social distancing

## Abstract

**Background:** Data on SARS-CoV-2 transmission from a pediatric index patient to others at the school setting are limited. Epidemiologic data on pediatric COVID-19 cases after school opening is warranted.

**Methods:** We analyzed data of the pediatric patients with COVID-19 collected from the press release of the Korea Centers for Disease Control and Prevention. Information on the school opening delay and re-opening policies were achieved from the press release from Korean Ministry of Education.

**Findings:** The school openings were delayed three times in March 2020. Online classes started from April 9, and off-line classes started from May 20 to June 8 at four steps in different grades of students. There was no sudden increase in pediatric cases after the school opening, and the proportion of pediatric cases remained around 7.0% to 7.1%. As of July 11, 45 children from 40 schools and kindergartens were diagnosed with COVID-19 after off-line classes started. More than 11,000 students and staff were tested; only one additional student was found to be infected in the same classroom. Among those 45, 32 (71.1%) patients had available information for the source of infection. Twenty-five (25/45, 55.6%) were infected by the family members. The proportions of pediatric patients without information on infection sources were higher in older age group (middle and high school students) than in younger age group (kindergarten and elementary school students) (47.6% *vs* 12.5%, *p*=0.010). In the younger age group, 79.1% of children were infected by family members, while only 28.6% of adolescents in the older age group were infected by family members (*p*<0.001).

**Interpretation:** Korea had a successful transition from school closure to re-opening with online and off-line classes. Although partial, off-line school opening did not cause significant school-related outbreak among pediatric population although young children and adolescents may have different epidemiologic features.

**Funding:** None.

## Introduction

Coronavirus disease 2019 (COVID-19) cases in the world are 12,041,795 as of July 11, 2020.^1^ Korea has about 51 million population, 18% of which is a pediatric population ≤19 years.^2^ The first COVID-19 patient in Korea was a Chinese tourist who was diagnosed on January 20, and the first pediatric patient was a 10-year Korean girl who was diagnosed on February 18, 2020.^3^ As of July 11, 2020, 964 Korean children ≤19 years and 214 children ≤9 years have been diagnosed. The proportions of pediatric cases ≤19 years and ≤9 years are 7.1% and 1.6% of all confirmed cases, respectively.^4^

Respiratory virus infection among school-aged children is an important epidemiologic consideration. Data on SARS-CoV-2 transmission from a paediatric index case to others at school settings are limited. A recent systematic review examined 31 studies on whether school closure had a quantifiable effect on influenza transmission and reported that school closure reduced the peak of the related outbreak by about 29.7% and delayed the peak by a median of 11 days.^5^ However, it is unclear whether school closures are effective in coronavirus outbreaks due to severe acute respiratory syndrome (SARS), Middle East respiratory syndrome (MERS), or especially SARS-CoV-2 of which transmission dynamics appear to be different.

It has been several months since schools are closed in more than 190 countries in the world, affecting 1.57 billion children, about 90% of the world student population. According to the United Nations Educational, Scientific and Cultural Organization (UNESCO), country-wide school closure peaked to 194 countries on April 13, 2020, and it decreased to 110 countries on July 11, 2020.^6^

Korea also delayed the school opening after winter vacation and closed the schools because of COVID-19. However, online and off-line school opening was proceeded by the government. We reviewed the pediatric epidemiology of COVID-19 in Korea according to the timeline of school opening delay and school re-opening using publicly available data.^7^ We examined whether school opening led the increase of pediatric COVID-19 cases in Korea, a country that had never been lockdown, only performed varying degrees of social distancing and rigorous contact tracing with fast testing whenever confirmed cases are identified.

## Methods

### Epidemiological investigation in Korea

Data of the confirmed patients with COVID-19 was collected from the press release by the Korea Centers for Disease Control and Prevention (KCDC) from February 18 (first pediatric patient diagnosed) to July 11, 2020.^4^ KCDC released the number of patients once or twice a day whenever new cases were identified until February 6. Then, they announced the numbers regularly twice a day from February 7 to March 1. From March 2, the number of cases summarized by 0 am on each day were announced the next day. Data for this report were collected from the announcement at 4 pm from February 18 to March 1, and at 0 am from March 2 to July 11.^4^

A confirmed case was defined as a person who was infected with SARS-CoV-2 regardless of clinical manifestations according to the diagnostic testing standard; genetic test (real-time RT-PCR) or virus isolation.^8^

### School opening delay and re-opening policy

Information on the school opening delay and re-opening policies was achieved from the press release by the Korean Ministry of Education (KMOE).^10,11^ Online classes were opened by using online education platforms such as “EBS Online class” of Korea Educational Broadcasting System (EBS), Digital Textbook Site, and E-learning Site.^9 11^ A decision of the off-line school opening was made in collaboration with KCDC, taking into account the opinions of school teachers, parents, and the Metropolitan and Provincial Offices of Education. After the decision, the guidelines were released for personal hygiene, virus prevention measures, procedures to follow when suspected symptoms occur in students.^10,11^ In addition, KMOE recommended that the number of students attending classes would not exceed a certain proportion to avoid overcrowding in schools. The specific method was determined by each school’s decision autonomously.^11^In the case of high school senior students (grade 12, G12), they attended the school for off-line classes daily considering the urgent academic need for the university entrance examination.^9^

Except for G12 students, the number of students who attended off-line classes was determined by the number of confirmed COVID-19 patients in the local area where the school is located and the characteristics of each school. 1) In cities or provinces where there were no confirmed cases recently, students of small schools (a total number of students of 60 or less) could attend school daily. The rest of the schools (Over 500-1,000 students) were recommended that the number of students attending school at the same time should not exceed two-thirds of the total students. 2) In areas where the confirmed cases were increasing, it was recommended that less than one-third of the total students attend the school at the same time. 3) When a confirmed case was identified at a school, the school would stop off-line classes and proceed to online classes during the investigation. Specific examples are listed in table 1.^9–11^

**Table 1.** Schedules and Method for School Attending According to the School Situation.

| **Frequency<sup>a</sup>** | **Distancing among students** | **School situation** |
| --- | --- | --- |
| Daily | Different school times<br>for each grade | Small schools or<br>no confirmed cases in that area |
| Once a week | One class per day<br>for each grade | Cities or provinces where the<br>confirmed cases are increasing<br>Elementary school (G1 to G6<br>that are relatively difficult to<br>perform personal hygiene) |
| Twice or three times a week | Two grades per day | 500 students or more<br>(G7 to G11, mainly) |
| Every other week | One grade per week | G10 and 11 in high school |

<sup>a</sup> Rest of the day, all students should attend the online class. G, grade.

At school, all students in the classroom should maintain social distancing. Upon arrival, students keep the distance before entering their classroom in the line (figure 1A). Teachers would check the body temperatures and monitor their symptoms. Schools are also preparing plastic barriers for lunchtimes, and hand sanitizers, and extra masks for additional emergency uses. In order to provide lunch safely, the contacts among students are minimized by keeping the distance in the waiting line (figure 1B) and arranging different lunchtimes for different grades.^7^ In the cafeteria, temporary plastic barriers are placed on the tables, and students eat in silence (figure 1C, 1D). Students and teachers wear masks at all time. However, students do not wear a mask at the playground as long as they keep the distance. Online classes are recommended for music classes to sing or play wind instruments.^9,10,14^

**Figure 1:**
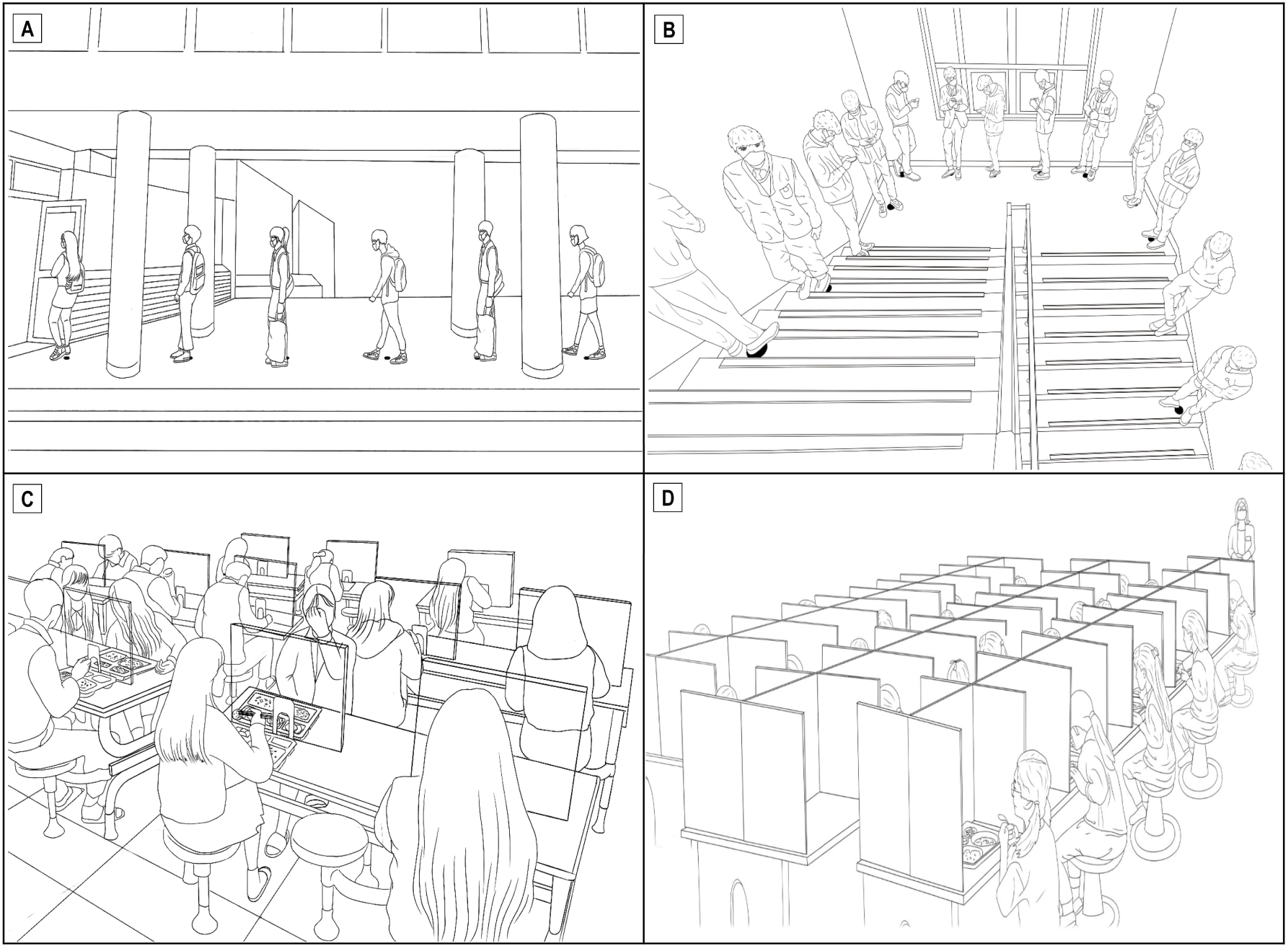
Illustrations showing how to avoid close contact after school opening. (A) Students are waiting with maintaining a distance to check their body temperature when they arrive at school. (B) High school students with masks are waiting in line, keeping a distance more than one meter from each other to enter the cafeteria. (C) High school students are sitting with space and eat lunch on a table with plastic barriers at the cafeteria. (D) Elementary school students sit with space and eat lunch on a table with plastic barriers at the cafeteria.

This study analyzed the data that is publicly available from the reports by KCDC and policy announcement by KMOE.^4,9^ Therefore, it was not considered that this study was subject to institutional review board approval.

## Results

### No lockdown but only social distancing strategies

The Korean government never closed the border to other countries or executed the lockdown strategy to control the outbreak of the nation even during the rapid surge in February and March. Instead, after raising the alert level from orange to red on February 23, it implemented varying degrees of social distancing strategies with rigorous contact tracing and massive and rapid testing on any suspicious cases.

Korean government executed social distancing (level II) on February 29 and further recommended enhanced social distancing (level III) on March 23. On April 20, with the relative control of the COVID-19 in the nation, the social distancing level was reduced from enhanced social distancing (level III) to social distancing (II). On May 6, dynamic distancing (level 1) has started (table 2).

**Table 2.** Mitigation Strategies and Dates of Implementation.

|  | Implementation date | Mitigation strategies | Degree of social distancing level |
| --- | --- | --- | --- |
| A | February 23 | Alert level raised to Red | - |
| B | February 29 | Social distancing | II |
| C | March 23 | Enhanced social distancing | III |
| D | April 20 | Social distancing | II |
| E | May 6 | Dynamic social distancing | I |
| F | May 21 | Administrative regulation of mass gathering | I |

Although the Korean government did not close the national border, various measures were implemented for international visitors or returning Koreans from aboard. For those who entered Korea from abroad, everybody was advised for self-isolation for 14 days from the entry and tested if symptomatic since April 1, 2020. From April 13, anybody who entered Korea from the U.S. or Europe was tested for SARS-CoV-2 regardless of symptoms within three days of arrival, and incoming travelers from other areas were tested when they had symptoms. From May 11, every inbound traveler should get tested within 14 days upon their arrival regardless of symptoms.

### School closure and opening delay according to the evolving stages of COVID-19 epidemiology in Korea

The first community transmission case without a travel history or an epidemiological link to previously confirmed cases was diagnosed on February 16 (29^th^ patient), about four weeks from the first patient of the nation reported on January 20. The heralding case for the epidemic surge was also identified on February 18 (31^st^ patient). This led the investigation of a religious group related massive outbreak in Daegu metropolitan area and Gyeongsangbuk-do (province) in February and Mach 2020, and a daily new case was recorded as high as 813 patients on February 29 (figure 2).

**Figure 2:**
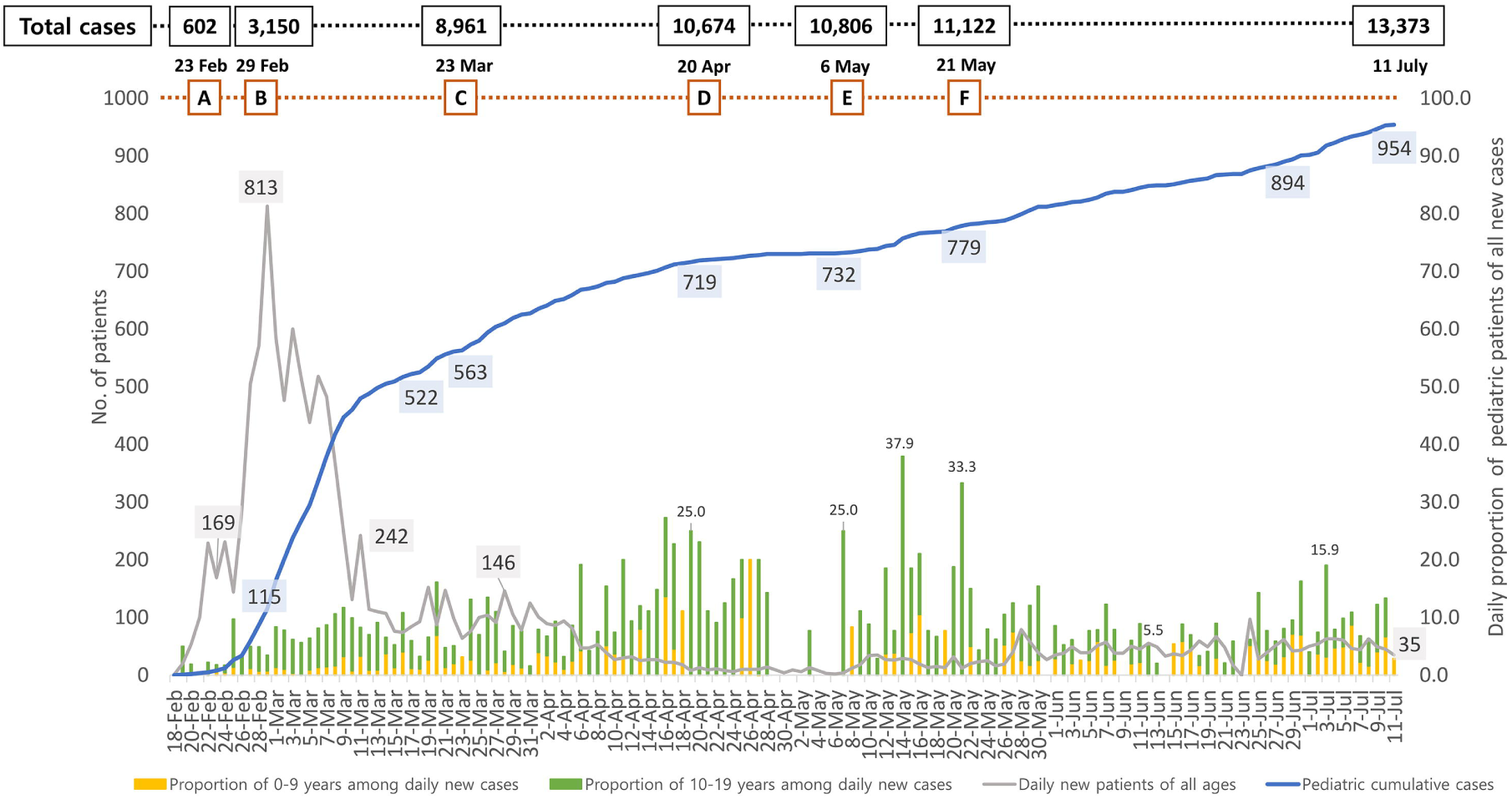
Epidemiology of pediatric patients with COVID-19 in Korea from February 18 to July 11, 2020. Levels of national alert and social distancing on different dates are shown in the alphabet (red squares), and the total numbers of confirmed patients on each date are also shown (black squares). The number of daily new cases of all ages (grey line), the cumulative number of pediatric cases (≤19 years old) (blue line), and the proportion of pediatric patients of all daily new cases (yellow and green bars) are shown.

The first pediatric COVID-19 patient was a 10-year old school-aged child and also diagnosed on February 18,^9^ and reported as the first pediatric case on February 19 by the KCDC formal briefing^4^. Since she was diagnosed during self-quarantine at home after the exposure to a family member, there was no school exposure because of this child. Figure 2 shows the cumulative number of pediatric cases from February 18 to July 11 and a proportion of pediatric patients of the daily new patients in all populations. It also shows the total number of COVID-19 cases on dates when the Korean government implemented a change for social distancing. During the surge of COVID-19 in Daegu metropolitan area and Gyeongsangbuk-do (province), the number of pediatric cases also rose abruptly as adult cases from one pediatric case on February 18 and 522 cases on March 17 when the third school opening delay was announced. Many of the older adolescents were also related to the religious group in those areas. Although the pediatric numbers also rose rapidly in February and early March, since the number of daily new patients was so high, the proportion of pediatric patients seemed relatively low.^12^

With this epidemic situation, the KMOE decided to delay the school opening for the new school year, which is usually set on March 2 every year. Eventually, the school openings were delayed three times in Korea (table 3). The first delay on school opening was decided when the national alert level was lifted from orange to red on February 23, because of the outbreak surge.

**Table 3.** Dates of School Opening Delay in Korea During COVID-19 Pandemic in 2020.

| Delay | Announcement day | Scheduled school opening dates | New opening dates |
| --- | --- | --- | --- |
| 1<sup>st</sup> | February 23 | March 02 | March 09 |
| 2<sup>nd</sup> | March 02 | March 09 | March 23 |
| 3<sup>rd</sup> | March 17 | March 23 | April 06 |

The second delay was announced on March 02 after the number of daily new patients recorded 813 on February 29 with the challenges in controlling the overwhelming outbreak in Daegu metropolitan area. The third delay was decided while further data were being sought, and safety measures were planned to help decision for school opening. In addition, the Korean government implemented enhanced social distancing (level III) since March 23, which was the scheduled date for the third school opening date. The numbers for daily new patients were still above 100 persons a day, and there were newly identified group outbreaks at the church near Seoul, the capital of the nation, and the nursing home in Daegu. Therefore, the school closure continued until Aril 06, 2020. Meanwhile, additional mathematical modelling and simulation study on school openings in Korea became available in early April 2020.^10^ Of note, the proportion of pediatric cases appeared high in mid-April and mid-May. However, the numbers of daily new cases of all populations were maintained at a low level (less than 50 cases per day), and the absolute number of new pediatric cases per day did not significantly increase.

### School opening and an impact on the epidemiology of COVID-19 in the pediatric population

The dates for school openings for students at different grades (online and off-line) are shown in table 4 and figure 3. The number of pediatric COVD-19 patients and the proportions of pediatric patients of all confirmed cases are shown in figures 3A and 3B with school opening dates.

**Table 4:** Step-wise School Opening Online and Off-line in Korea During COVID-19 Pandemic in 2020.

| Online school opening | Dates | School Grades (G) |
| --- | --- | --- |
| 1<sup>st</sup> | April 9 | G12, G9 |
| 2<sup>nd</sup> | April 16 | G11-10, G8-7, G6-4 |
| 3<sup>rd</sup> | April 20 | G3-1 |
| Off-line school opening | Dates | School Grades (G) |
| 1<sup>st</sup> | May 20 | G12 |
| 2<sup>nd</sup> | May 27 | G11, G9, G2-1, Kindergarten |
| 3<sup>rd</sup> | June 3 | G10, G8, G4-3 |
| 4<sup>th</sup> | June 8 | G7, G6-5 |

In Korean education system, children in grades 1-6 are in elementary schools, students in grades 7-9 in middle schools and students in grades 10-12 are in high schools.

**Figure 3:**
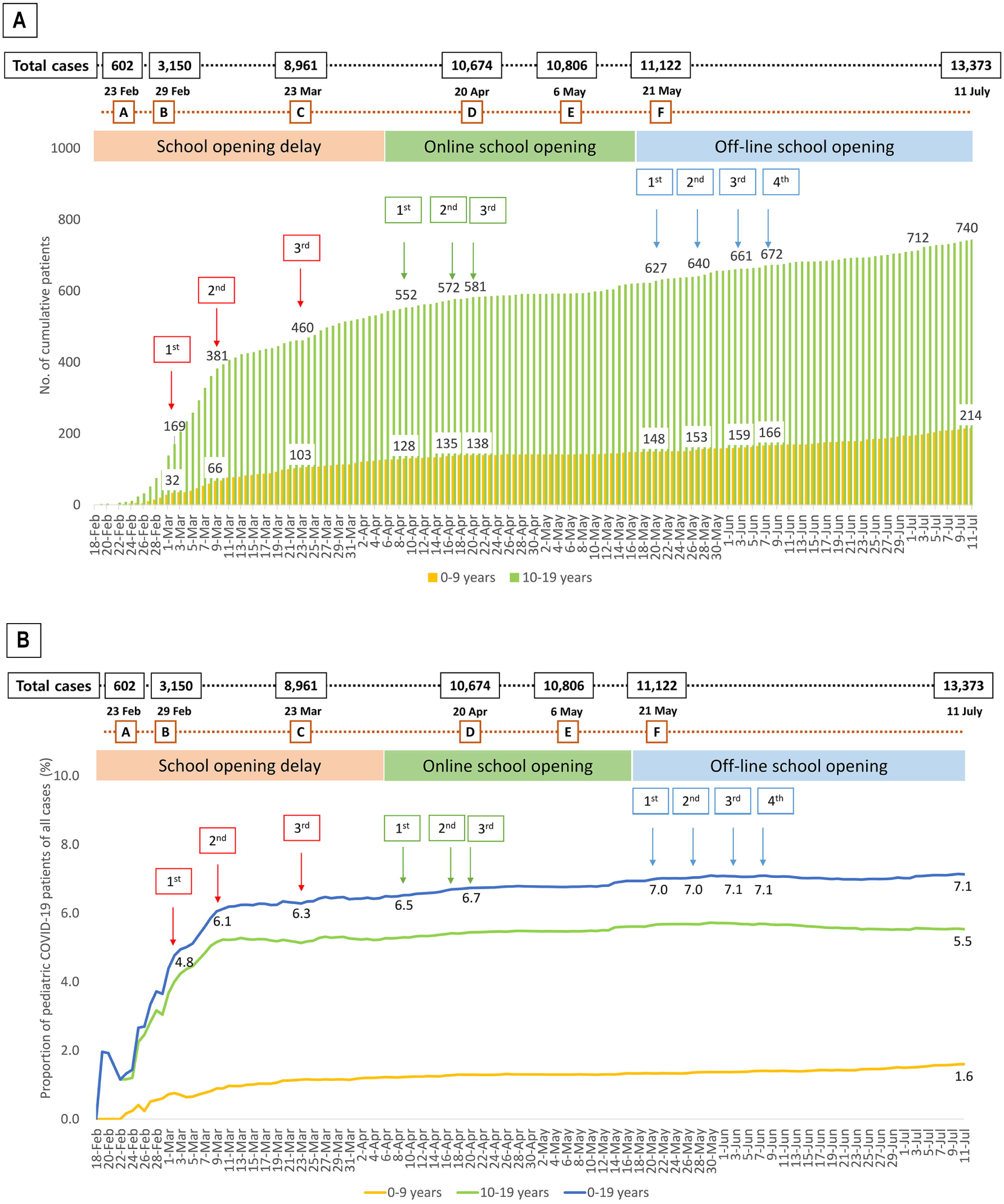
(A) Number of cumulative pediatric patients (0-9 years and 10-19 years) with COVID-19 and school opening delay and re-opening dates. (B) Proportions of cumulative pediatric patients (0-19 years) with COVID-19 and school opening delay and re-opening dates. Levels of national alert and social distancing on different dates are shown in the alphabet (red squares), and the total numbers of confirmed patients on each date are also shown (black squares).

Online classes started at three steps on April 9, April 16, and April 20, with high school senior students (12^th^ grade, G12) and middle school senior students (9^th^ grade, G9) first. Then, younger students started online classes. As of April 9, when the first online classes were open, the number of pediatric COVID-19 cases (≤19 years) was 680, and the proportion of pediatric patients of all confirmed cases was 6.5%.

Off-line classes started at four steps on May 20, May 27, June 3, and June 8 with high school senior students (12^th^ grade, G12) who went to school first in the 2020 school year. As of May 20, when the first off-line classes were open, the number of pediatric COVID-19 cases (≤19 years) was 775, and the proportion of pediatric patients of all confirmed cases was 7.0%. As of July 11, 52 days from the first off-line opening, the number of pediatric COVID-19 cases (≤19 years) was 954, and the proportion of pediatric patients of all confirmed cases was 7.1%.^4^

There was no obvious trend for the sudden increase of pediatric cases or the proportion of pediatric cases of all confirmed cases in the nation before and after the school opening. ^5^

### SARS-CoV-2 exposure at school setting after off-line opening

Table 5 and Figure 4 summarized SARS-CoV-2 exposures and investigation at the school settings where a pediatric case was the index case since the off-line school opening on May 20. As of July 11, 45 children (index students) attended 40 schools and kindergartens and caused exposures in other students and staff members: twelve high schools, eight middle schools, fifteen elementary schools, and five kindergartens. Among 45 pediatric cases, 32 (71.1%) students were available for the source of infection, twenty-five (78.1%, of cases with available source, 55.6% of total cases) students were infected by their family members (figure 4). The two students were from the church outbreak, and five students were infected from the academy outbreak. ^4,9,13,14,10,16–18^

**Table 5.** Summary of SARS-CoV-2 exposures by index students at schools and kindergarten after school re-opening.

|  | **Kindergarten**<br>**(4-5 years)** | **Elementary school**<br>**(6-12 years)** | **Middle school**<br>**(13-15 years)** | **High school**<br>**(16-18 years)** | **Total** |
| --- | --- | --- | --- | --- | --- |
| **Number of students** | 5 | 19 | 8 | 13 | 45 |
| **Known infection source N, (%)** | 4 (80.0) | 17 (89.5) | 5 (62.5) | 6 (46.2) | 32 (71.1) |
| **House hold infection N, (%)** | 4 (80.0) | 15 (78.9) | 5 (62.5) | 1 (7.7) | 25 (55.6) |
| **Unknown infection Source N, (%)** | 1 (20.0) | 2 (10.5) | 3 (37.5) | 7 (53.8) | 13 (28.9) |
| **Number of exposed schools** | 5 | 15 | 8 | 12 | 40 |
| **Tested individuals (N)** | ≥670 | ≥ 2,453 | ≥ 1,962 | ≥ 4,747 | ≥ 10,903 |
| **Exposed to two brothers (N)** | - | 1,071* |  | - |  |
| **Secondary infection (N)** | 0 | 1 | 0 | 0 | 1 |

\* Additional 1,071 individuals tested because of exposures from two brothers in the same family, one was an elementary school student, and the other was a middle school student.

**Figure 4:**
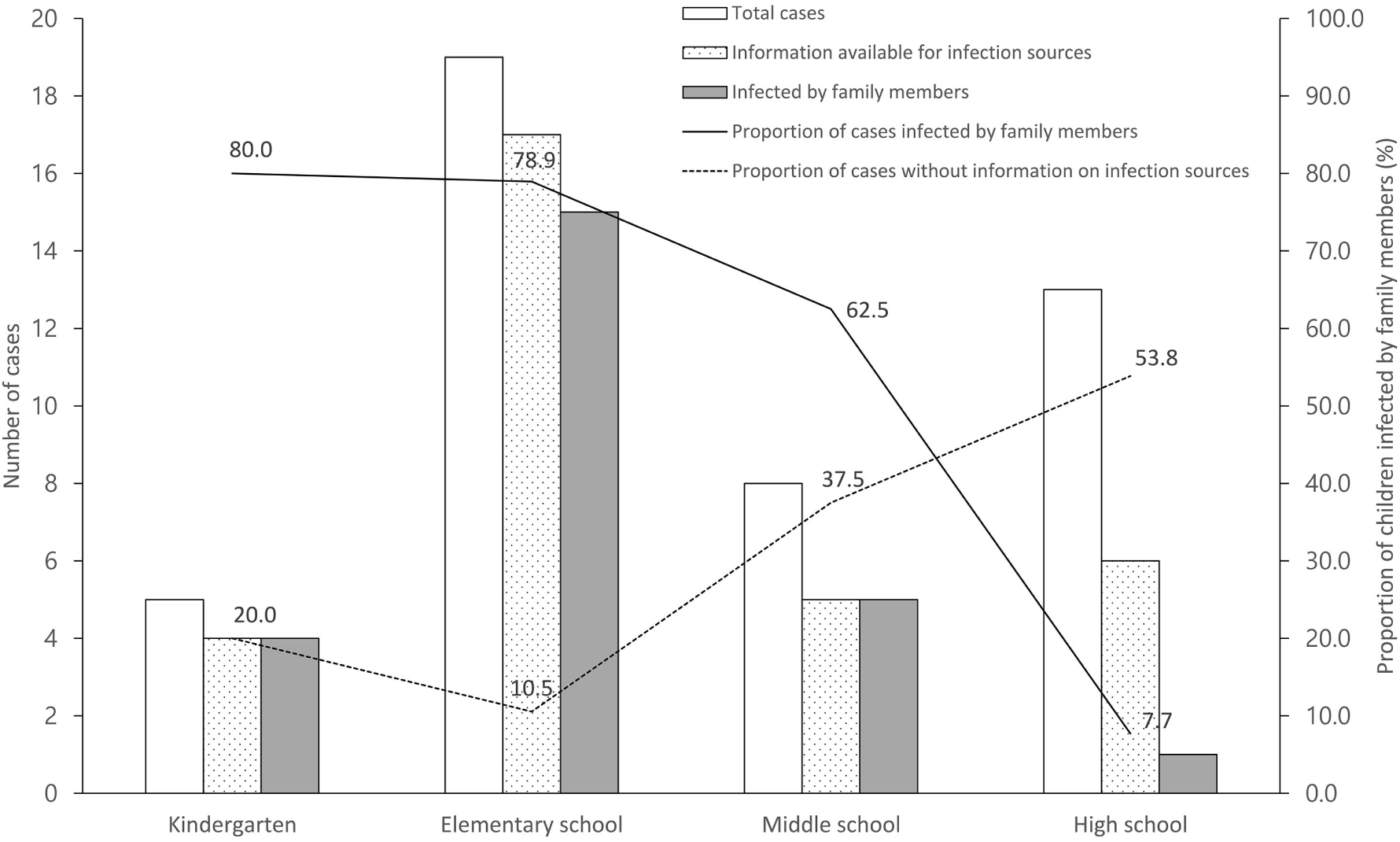
Number of paediatric patients in each school setting and information availability for infection sources.

At kindergartens, out of five children, four children (80%) were infected by their family members, and the information on the source of infection was not available in one child. More than 600 children and staff members in the kindergartens were tested for SARS-CoV-2. There were no secondary cases from kindergarten exposures.^11,14,16^

At elementary schools (G1-G6, 7-12 years), 19 cases were confirmed, and more than 3,000 students and staff members were tested. Among 19 cases, the source of infection was available in 17 (89.5%) cases, and 15 students were infected by the family members (78.9%). There was only one elementary school where the secondary cases were observed; one 11-year-old child in 5th grade transmitted the virus to two other children. One child was infected in the same classroom, and the other was not in the same class but was infected at the same exercising gym out of school. ^4,9,15^

At middle school (G7-G9, 13-15 years) and high schools (G10-G12, 16-18 years), 21 students were confirmed, and more than 7,500 students and staff members were tested. In middle school exposures, among eight adolescents, five were infected by family members (5/8, 62.5%). In high schools, only one adolescent was infected by family members (1/13, 7.7%). There were no secondary cases from middle school and high school exposures (Table 5). Of note, five students (G12) from four different high schools were infected at the same acting academy in Daegu. In the academy, they practised acting and singing for their university entrance examination without wearing a mask. ^14^ However, they did not transmit SARS-CoV-2 in their own school where they followed the rules for distancing among students and wearing a mask.^4,9^

The proportions of index students who did not have information on infection sources were higher in older age group (middle school and high school students) than in younger age group (kindergarten and elementary school students) (47.6% *vs* 12.5%, p=0.010). In the younger age group, almost 80% of children were infected by family members, but the proportion of students infected by family members decreased with age (p<0.001) (Figure 4).

## Discussion

This report describes the impact of school opening on pediatric COVID-19 epidemiology in Korea, a country that never closed its country border, or implemented country lockdown but only performed various degrees of social distancing strategies. There was no significant increase in the number of pediatric patients or the proportion of pediatric patients among all confirmed cases in the nation after the off-line school opening. In addition, there were no explosive secondary cases after school exposure by the index students at schools.

Closing and re-opening of schools and preschools is a major educational, political, and public health issue worldwide. The concerns about the transmission of the virus among children and teenagers in closed space of school have been a major drawback for the school opening decisions. However, the negative impact of limiting social and physical activities for an extended period of time in children is worrisome. In addition, significant inequalities among children with a high- and low-socioeconomic environment may exist in online learning accessibility and the level of care provided at home with prolonged school closure.

It is well-known that children are susceptible to many respiratory virus infections with frequent epidemics in the communities. Therefore, from the beginning of the COVID-19 outbreak, there were concerns about the role of pediatric population for the spread of the SARS-CoV-2. However, early transmission clusters were identified mainly from adults, making children less likely to be index cases in households or the communities. As data accumulated, a significant proportion of pediatric patients are known to be asymptomatic or mildly symptomatic^12,16^ with an exception for unusual cases with multisystem inflammatory syndrome in children (MIS-C).^17^ In addition, pediatric patients with COVID-19 also appeared to have similarly high viral loads at the early stage of illness as in adult patients.^18–20^ Therefore, the possibility for virus transmission from pre-symptomatic or mildly symptomatic pediatric index case(s) to others can also be questioned.^20,21^ However, there are little data that reported the transmission among children that led to a massive outbreak, especially in school exposure. A systematic review also suggested that children seem unlikely to be the main drivers of the COVID-19 pandemic.^22^ It is not clear whether the viral transmission dynamic in the pediatric population is different from that of adults, or this may be simply because most schools are closed for extended periods of time. There may be certain different features in pediatric population that need to be further explored.

It is of interest that of more than 11,000 students and staffs tested from 40 schools and preschools, only one additional student was infected in the same classroom at the elementary school. Based on the data summarized in this study, we consider that spread of COVID-19 within Korean schools has been very limited even after off-line classes started while following the rules for symptom monitoring and distancing among students. Since rigorous contact tracing and tests are being performed in Korea from the early phase of pandemic, there is little possibility that there are hidden pediatric cases. In addition, a recent seroepidemiological survey reported that only 0.03% of tested positive for SARS-CoV-2 antibody among 3,055 Koreans.^23^ Therefore, we consider that what we observed in this study would represent the real situation of the Korean pediatric population.

It is of note that the infection sources were more likely available in younger age group (kindergarten and elementary school) than the older age group. When the information on infection source was available, the majority of the young children (kindergarten and elementary students) were infected by family members. In a study from China, they also reported that 90.1% of 171 pediatric cases were related to family clusters.^24^ In a study from Switzerland on 40 pediatric cases, they investigated the dynamics of infection in their families; one or more adult family member was suspected or confirmed for COVID-19 before symptom onset of the study child in 79% of households.^25^ Therefore, the most common infection sources for pediatric cases appear to be their adult family members; children are not the primary source of infection or did not cause large outbreaks at school.^22,26^ However, careful attention should be paid to older adolescents who may have more sources of infection outside of the household.

The results of this report need to be interpreted with caution because even though the schools in Korea opened for off-line classes, it is a partial opening with significant measures to follow and restrictions. Therefore, this may not represent the full-day, five days a week school opening situation and may not apply to other countries where public control measures and test capacity differ. However, Korean experience on opening schools during this COVID-19 pandemic will provide some hopes that the students can go back to schools for off-line classes as long as they follow the safety measures for careful symptom monitoring, personal hygiene, wearing masks, and distancing among students that are known to reduce the transmission risk.^27^

Although there are several reports from the school exposure investigation in various countries.^28,29^ There has been no report that systematically described national progress on school closure, online opening, and off-line opening with an epidemiological analysis of the national pediatric population.

On March 10, 2020, the United Nations Children’s Fund (UNICEF), the International Federation of the Red Cross, and the WHO issued a guidance document on re-opening schools.^30^ The guidance considers the balance of risks to children’s health, well-being, learning, and development posed by disease transmission versus not attending school.

In conclusion, we observed that Korea had a successful transition from school closure to re-opening for online and off-line classes with various efforts to keep our pediatric population safe while they attend the school during COVID-19 pandemic. As this pandemic continues, we need to share our wisdom and scientific evidence for proper strategies to open schools safely in each community.

## Contributors

Yoonsun Yoon, Kyung-Ran Kim, and Hwanhee Park designed the study and data collection methods, did the initial data analyses, drafted the manuscript, and approved the final manuscript as submitted. Soyoung Kim contributed to the study concept and design and critically reviewed the manuscript. Yae-Jean Kim conceptualized and designed the study and critically reviewed and revised the manuscript. All authors approved the final submitted manuscript.

## Data Availability

Data of the confirmed patients with COVID-19 was collected from the press release by the Korea Centers for Disease Control and Prevention. Information on the school opening delay and re-opening policies was achieved from the press release by the Korean Ministry of Education.

## Acknowledgements

We appreciate the cooperation of the children, their parents, school staff, and public health officials at health care centers, the medical staff of the healthcare facilities of Korea. We thank the KCDC and KMOE for their efforts in responding to COVID-19 outbreak and school-related policies.

## References

1 COVID-19 Dashboard by the Center for Systems Science and Engineering (CSSE) at Johns Hopkins Uiversity (JHU), JOHNS HOPKINS CORONAVIRUS RESOURCE CENTER. https://coronavirus.jhu.edu/map.html. (accessed July 11. 2020.)

2 Korean Statistical Information Service. 2018 population census updated by the KOSIS, August 29, 2019 (Updated 2020). http://kosis.kr/statisticsList/statisticsListIndex.do?menuId=M_01_01&vwcd=MT_ZTITLE&parmTabId=M_01_01#SelectStatsBoxDiv (accessed July 16, 2020).

3 Korean Society of Infectious Diseases, Korean Society of Pediatric Infectious Diseases, Korean Society of Epidemiology, et al. Report on the Epidemiological Features of Coronavirus Disease 2019 (COVID-19) Outbreak in the Republic of Korea from January 19 to March 2, 2020. J Korean Med Sci 2020; 35(10): e112.

4 Daily Cumulative Confirmed Data, March 1 to July 11, 2020, Korea Centers for Disease Control and Prevention, Korea. 2020. http://www.cdc.go.kr/.

5 Bin Nafisah S, Alamery AH, Al Nafesa A, Aleid B, Brazanji NA. School closure during novel influenza: A systematic review. J Infect Public Health 2018; 11(5): 657–61.

6 UNESCO. COVID-19 impact on education as of July 11. https://en.unesco.org/covid19/educationresponse (accessed July 16, 2020).

7 Ministry of Education, Korea. Student health information center. http://www.schoolhealth.kr/web/bbs/selectNewBBSList.do?lstnum1=3041 (accessed July 11, 2020).

8 Korea Centers for Disease Control and Prevention. Coronavirus disease-19, Case definition. http://ncov.mohw.go.kr/en/baroView.do?brdId=11&brdGubun=112&dataGubun=&ncvContSeq=&contSeq=&board_id=&gubun=(accessed July 11, 2020).

9 Press release. Korean Ministry of Education, Sejong 2020. http://english.moe.go.kr/boardCnts/list.do?boardID=265&m=0301&s=english(accessed July 11 2020).

10 Press release. Seoul Metropolitan Office of Education, Seoul 2020. http://enews.sen.go.kr/news/list.do?step1=3&step2=1(accessed July 11 2020).

11 Notice for off-line class. Daegu Metropolitan Office of Education, Daegu 2020. http://www.dge.go.kr/main/cm/cntnts/cntntsView.do?mi=5652&cntntsId=32712020(accessed July 11 2020).

12 Choi SH, Kim HW, Kang JM, Kim DH, Cho EY. Epidemiology and clinical features of coronavirus disease 2019 in children. Clin Exp Pediatr 2020; 63(4): 125–32.

13 COVID-19 situation of Gwangju, Gwangju Metropolitan City. 2020. https://www.gwangju.go.kr/c19/c19/contentsView.do?pageId=coronagj2 (accessed July 11, 2020)

14 COVID-19 situation, updated July 11, 2020, Daegue Metropolitan Office OfEducation, Daegu. 2020. http://www.dge.go.kr/main/na/ntt/selectNttInfo.do?nttSn=1744944&mi=52882020). (accessed July 11, 2020)

15 COVID-19 situation of Daejeon, update July 11, 2020. Daejeon Metropolitan City. 2020. <https://www.daejeon.go.kr/corona19/index.do (accessed July 11, 2020).

16 Wu Z, McGoogan JM. Characteristics of and Important Lessons From the Coronavirus Disease 2019 (COVID-19) Outbreak in China: Summary of a Report of 72314 Cases From the Chinese Center for Disease Control and Prevention. JAMA 2020; 323(13): 1239–42.

17 Whittaker E, Bamford A, Kenny J, et al. Clinical Characteristics of 58 Children With a Pediatric Inflammatory Multisystem Syndrome Temporally Associated With SARS-CoV-2. JAMA 2020.

18 Han MS, Seong MW, Kim N, et al. Viral RNA Load in Mildly Symptomatic and Asymptomatic Children with COVID-19, Seoul. Emerg Infect Dis 2020; 26(10).

19 Wolfel R, Corman VM, Guggemos W, et al. Virological assessment of hospitalized patients with COVID-2019. Nature 2020; 581(7809): 465–9.

20 Jones TC, Mühlemann B, Veith T, et al. An analysis of SARS-CoV-2 viral load by patient age. medRxiv 2020.

21 He X, Lau EHY, Wu P, et al. Temporal dynamics in viral shedding and transmissibility of COVID-19. Nat Med 2020; 26(5): 672–5.

22 Ludvigsson JF. Children are unlikely to be the main drivers of the COVID-19 pandemic - A systematic review. Acta Paediatr 2020; 109(8): 1525–30.

23 Korea Centers for Disease Control and Prevention. Updates on COVID-19 in Republic of Korea (as of 9 July, 2020), Korea. http://ncov.mohw.go.kr/tcmBoardView.do?brdId=&brdGubun=&dataGubun=&ncvContSeq=355336&contSeq=355336&board_id=&gubun=ALL (accessed July 11, 2020).

24 Lu X, Zhang L, Du H, et al. SARS-CoV-2 Infection in Children. N Engl J Med 2020; 382(17): 1663–5.

25 Posfay-Barbe KM, Wagner N, Gauthey M, et al. COVID-19 in Children and the Dynamics of Infection in Families. Pediatrics 2020.

26 Lee B, Raszka WV, Jr. COVID-19 Transmission and Children: The Child Is Not to Blame. Pediatrics 2020.

27 Chu DK, Akl EA, Duda S, et al. Physical distancing, face masks, and eye protection to prevent person-to-person transmission of SARS-CoV-2 and COVID-19: a systematic review and meta-analysis. The Lancet 2020.

28 National Centre for Immunisation Research and Surveillance (NCIRS), Australia. COVID-19 in schools – the experience in NSW. http://ncirs.org.au/sites/default/files/202004/NCIRS%20NSW%20Schools%20COVID_Summary_FINAL%20public_26%20April%202020.pdf (accessed July 11, 2020).

29 Heavey L, Casey G, Kelly C, Kelly D, McDarby G. No evidence of secondary transmission of COVID-19 from children attending school in Ireland, 2020. Euro Surveill 2020; 25(21): 2000903.

30 WHO. COVID-19. IFRC, UNICEF and WHO issue guidance to protect children and support safe school operations. https://www.who.int/news-room/detail/10-03-2020 (accessed May 26, 2020).

